# Interplay between Chronic Kidney disease, Hypertension, and Stroke: Insights from a Multivariable Mendelian Randomization Analysis

**DOI:** 10.1101/2022.09.14.22279923

**Authors:** Dearbhla M. Kelly, Marios K. Georgakis, Nora Franceschini, Deborah Blacker, Anand Viswanathan, Christopher D. Anderson

## Abstract

**Background and Objectives:** Chronic kidney disease (CKD) increases the risk of stroke, but the extent through which this association is mediated by hypertension is unknown. We leveraged large-scale genetic data to explore causal relationships between CKD, hypertension and cerebrovascular disease phenotypes.

**Methods:** We used data from genome-wide association studies (GWAS) of European ancestry to identify genetic proxies for kidney function (CKD diagnosis, estimated glomerular filtration rate [eGFR], and urinary albumin-to-creatinine ratio [UACR]), systolic blood pressure (SBP), and cerebrovascular disease (ischaemic stroke and its subtypes, and intracerebral haemorrhage [ICH). We then conducted univariable, multivariable and mediation Mendelian randomization (MR) analyses to investigate the effect of kidney function on stroke risk and the proportion of this effect mediated through hypertension.

**Results:** Univariable Mendelian randomization revealed associations between genetically determined lower eGFR and risk of all stroke (OR per 1-log decrement in eGFR, 1.77; 95% CI, 1.31-2.40; p<0.001), ischaemic stroke (OR, 1.81; 95% CI, 1.31-2.51; p<0.001), and most strongly with large artery stroke (LAS) (OR, 3.00; 95% CI, 1.33-6.75; p=0.008). These associations remained significant in the multivariable MR analysis, controlling for SBP (OR, 1.98; 95% CI, 1.39-2.82; p<0.001 for AS; OR, 2.16; 95% CI, 1.48-3.17; p<0.001 for IS; OR, 4.35; 95% CI, 1.84-10.27; p=0.001 for LAS). with only a small proportion of the total effects mediated by SBP (10.5%, 6.6% and 7.8%, respectively). Total, direct and indirect effect estimates were similar across a number of sensitivity analyses.

**Discussion:** Our results demonstrate an independent causal effect of impaired kidney function, as assessed by decreased eGFR, on stroke risk, particularly LAS, even when controlled for SBP. Targeted prevention of kidney disease could lower atherosclerotic stroke risk independent of hypertension.

## INTRODUCTION

Chronic kidney disease (CKD) carries a strong epidemiologic association with stroke risk.^1, 2^ In a large meta-analysis of 83 studies, the risk of stroke increased by 7% for every 10ml/min/1.73m^2^ decrease in estimated glomerular filtration rate (eGFR) and by 10% for every 25 mg/mmol increase in urine albumin–creatinine ratio (UACR).^3^ However, the central mechanisms underpinning the relationship between CKD and stroke risk remain unclear.^4^ Traditional risk factors such as hypertension, diabetes and atrial fibrillation (AF) are all highly prevalent in patients with CKD but non-traditional CKD-related risk factors such as chronic inflammation, uremic toxins, and mineral-bone disorder are also purported to contribute to risk by triggering vascular injury and endothelial dysfunction.^5^

Because hypertension is comorbid in up to 90% of patients with CKD,^6^ it is not clear whether the relationship between CKD and stroke is truly independent of blood pressure (BP). We previously performed two systematic reviews and meta-analysis of low eGFR, proteinuria and stroke risk, analysing studies according to the way in which they adjusted for hypertension.^7, 8^ For pooled studies of low eGFR and stroke risk, there was near-complete attenuation of associations after adjustment for longitudinal BP control.^7^ In contrast, in the meta-analysis of proteinuria, the pooled risk association did not substantially attenuate even with the extensive adjustment for BP.^8^

Mendelian randomization (MR) is a method that uses genetic variants associated with modifiable exposures as instrumental variables to test the causal effect of a risk factor on a disease or health-related outcome.^9^ A prior univariable MR study found associations between genetically determined lower eGFR and higher UACR with increased risk of large artery stroke (LAS), as well as between genetically determined higher UACR with increased risk of intracerebral haemorrhage (ICH).^10^ Building on this work, multivariable MR (MVMR) offers the opportunity to further explore the role of hypertension as a potential mediator in the relationship between CKD and stroke. MVMR uses genetic variants associated with multiple, potentially related exposures to estimate the effect of each on a single outcome.^11^ It also allows for estimation of mediation effects.

Therefore, using MVMR, we leveraged large-scale genetic data to explore causal relationships between genetic predispositions to CKD, hypertension and cerebrovascular disease phenotypes, testing the hypothesis that the effect of CKD on stroke risk is mediated through increased BP.

## METHODS

### Study Design

We conducted summary-level univariable, multivariable and mediation MR analyses to investigate the effect of kidney function on stroke risk and the proportion of this effect mediated through hypertension. In MR analysis, a genetic variant serves as a valid instrument if certain assumptions hold, including that the genetic variant is associated with the exposure of interest, there are no common causes of genotype and health outcomes, and the genetic variant affects the outcome only through their effect on the risk factor of interest.^9^ *Figure 1* conceptually depicts our approach, and *Table 1* summarizes the data sources used in each analysis.

**Figure 1:**
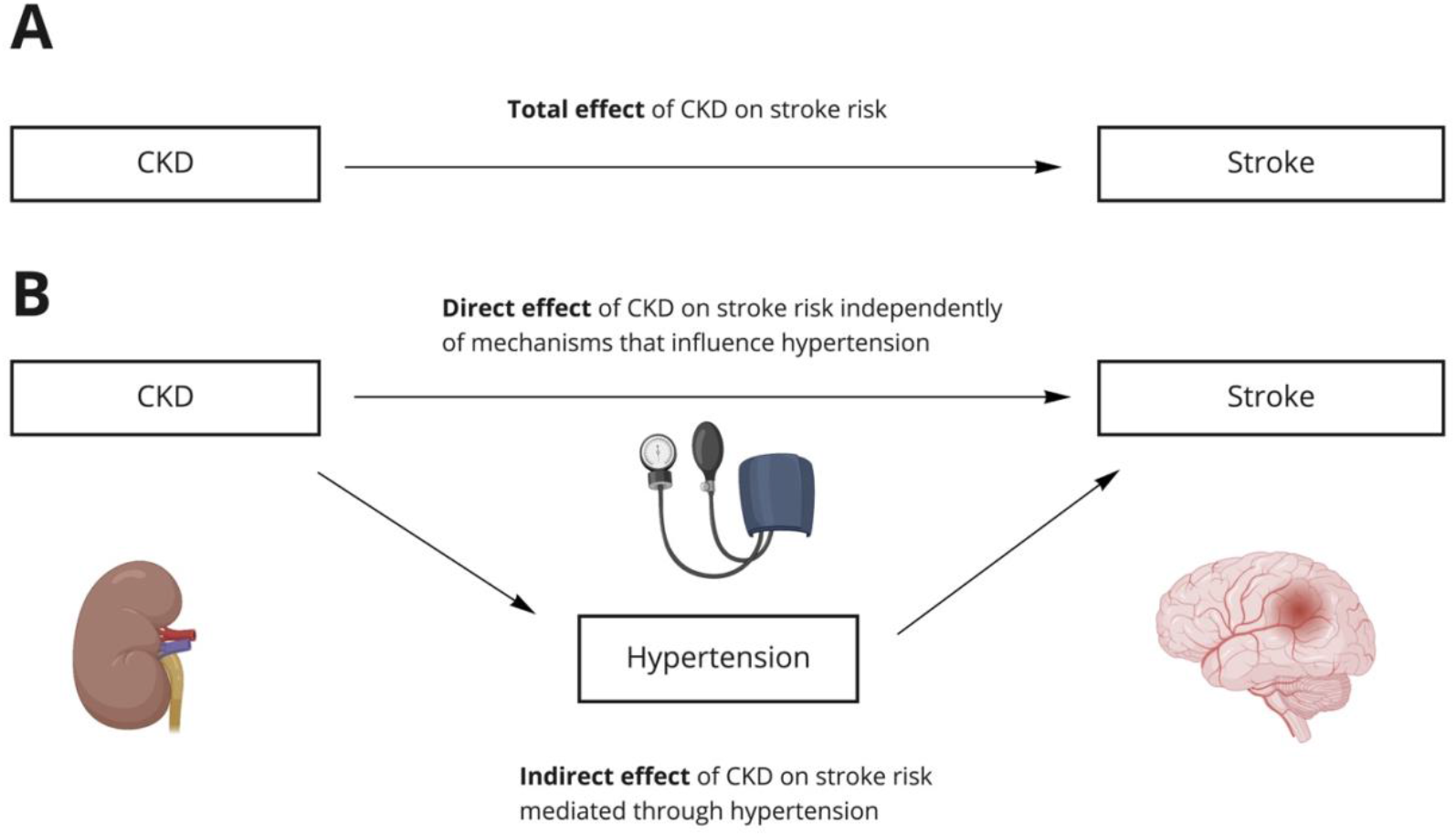
Direct acyclic graph to illustrate total, direct, and indirect effects of chronic kidney disease (CKD) on stroke risk. Directed acyclic graphs demonstrating the hypothesized direction for the total effect of CKD on increased odds of stroke (A) and the hypothesized direction for the effect of CKD on increased hypertension (B), which may partially mediate the effect of CKD on stroke risk.

**Table 1:** Characteristics of participants included in the genome-wide association studies utilised for the analyses.

| <u>Traits</u> | <u>Consortium</u> | <u>Participants, n</u> | <u>Women, %</u> | <u>Mean age, y (SD)</u> | <u>Trait mean value (SD)</u> | <u>GWAS adjustments</u> | <u>Imputation panel used</u> |
| --- | --- | --- | --- | --- | --- | --- | --- |
| **Kidney traits** |  |  |  |  |  |  |  |
| **CKD** | CKDGen | 480,698<br>(41,395 cases & 439,303 controls) | 50.0 | 54.0 |  | Age, sex. | Haplotype Reference Consortium v1.1 or 1000 Genomes Project phase 3 v5 (1000Gp3v5) ALL or phase 1 v3 (1000Gp1v3) ALL panel. |
| **eGFR** |  | 460,826 | 54.0 | 56.8 (8.0) | 88.1 (16.1) ml/min/1.73m<sup>2</sup> | Age, sex and other study-specific covariates. | Haplotype Reference Consortium (HRC, v1.1) or the 1000 Genomes Project (ALL panel) panels. |
| **UACR** | UK Biobank | 382,500 | 54.0 | 56.9 (8.3) | 9.8 (2.7) mg/g | Age, sex, and the first ten genetic PCs | Haplotype Reference Consortium reference panel |
| **Cerebrovascular disease phenotypes** |  |  |  |  |  |  |  |
| **AS** | MEGASTROKE | 40,585 cases & 406,111 controls | 41.7 | 67.4 (12.3) |  | Age, sex. | All studies used imputed genotypes based on at least the 1000G phase 1 multi-ancestral reference panel |
| **IS** |  | 34,217 cases & 406,111 controls | 41.7 | 67.4 (12.3) |  |  |  |
| **LAS** |  | 4,373 cases & 406,111 controls | 48.9 | 65.9 (10.4) |  |  |  |
| **SVS** |  | 5,386 cases & 406,111 controls | 45.5 | 65.6 (12.4) |  |  |  |
| **CES** |  | 7,193 cases & 406,111 controls | 46.4 | 68.1 (9.4) |  |  |  |
| **ICH** | ISGC | 1,545 cases & 1,481 controls | 45.1 | 67.0 (10.0) |  |  |  |
| **Blood pressure** |  |  |  |  |  |  |  |
| **SBP** | UK Biobank/ICBP | 757,601 | 52.6 | 55.63 (8.9) | 139 (20) | Age, sex, age2, BMI and a binary indicator variable for UKB vs UK BiLEVE to account for the different genotyping chips. | For UKB - Haplotype Reference Consortium (HRC) panel. For ICBP - either the 1000 Genomes Project Phase 1 integrated release v.3 [March 2012] all ancestry reference panel<br>or the HRC panel |
| **DBP** |  | 757,599 |  |  |  |  |  |
| **Carotid traits** |  |  |  |  |  |  |  |
| **CIMT** | CHARGE | 71,128 |  |  |  | Age, sex and<br>up to 10<br>principal<br>components. | Each GWAS<br>study conducted<br>genome-wide<br>imputation using<br>a Phase 1<br>integrated<br>reference panel<br>from the 1000G<br>Consortium<br>using IMPUTE2<br>or<br>MaCH/minimac,<br>and used Human<br>Reference<br>Genome Build<br>37. |
| **Carotid plaque** |  | 48,434 |  |  |  |  |  |

To further explore associations between genetically predicted kidney disease traits and LAS, we additionally performed Mendelian randomization analyses to investigate the relationship between lower eGFR, carotid intima media thickness (CIMT) and the presence of carotid plaque.

### Data Sources

We used data from GWAS of European ancestry to identify genetic proxies for kidney function (CKD diagnosis, eGFR, and UACR), blood pressure (systolic blood pressure [SBP], diastolic blood pressure [DBP]) and cerebrovascular disease (ischaemic stroke and its subtypes, and ICH). Similar to previous work,^10^ we used CKD diagnosis as well as both eGFR and UACR to explore impaired kidney function since these exposures may capture different aspects of kidney pathophysiology and their combined assessment of function is increasingly recommended in both clinical practice and research.^12, 13^ The cystatin-C-based formula for estimating eGFR was chosen instead of creatinine-based ones due to the higher accuracy and better cardiovascular predictive ability of the former.^14, 15^

CKD diagnosis and eGFR GWAS summary meta-analysis statistics were obtained from the latest 1000 Genomes-based Chronic Kidney Disease Genetics (CKDGen) consortium effort (ckdgen.imbi.uni-freiburg.de/). The CKDGen consortium with the correspondent genotypic and phenotypic assessment procedures that led to the GWAS results, are described elsewhere.^16, 17^ Briefly, genetic instrumental variables for eGFR were identified from a GWAS meta-analysis from the CKDGen Consortium and UK Biobank (n = 460,826 for cystatin-based eGFR).^17^ For CKD diagnosis, we obtained genetic association estimates from an earlier CKDGen GWAS meta-analysis (n=480,698 [41,395 cases and 439,303 controls]).^16^ GWAS results for the UACR trait were derived from the latest study that leveraged the UK Biobank data (n=382,500).^18^

Association estimates for cerebrovascular phenotypes were derived from the MEGASTROKE consortium^19^ and the International Stroke Genetic Consortium (ISGC) GWAS for ICH.^20^ Detailed descriptions of study populations and stroke subtyping ascertainment are available (cerebrovascularportal.org/). Briefly, we used the GWAS summary statistics of the European ancestry analysis of the study (40,585 cases; 406,111 controls). The phenotypes used (1) All stroke (AS), inclusive of all subtypes; (2) Ischaemic stroke (IS) regardless of subtype; the 3 available aetiologic ischaemic subtypes (3) Large artery stroke (LAS), (4) Small vessel stroke (SVS), and (5) Cardioembolic stroke (CES); and (6) ICH. IS and ICH were defined based on clinical and imaging criteria, whereas IS subtypes were based on the Trial of Org 10172 in Acute Stroke Treatment (TOAST) classification system.^21^

SBP and DBP summary meta-analysis statistics were obtained from the largest genetic association study of blood pressure traits to-date which combines data from UK Biobank and the International Consortium of Blood Pressure (grasp.nhlbi.nih.gov/FullResults.aspx).^22^ The study included 757,601 individuals of European ancestry. Briefly, SBP values were based on the average of 2 values and adjusted for antihypertensive therapy.

CIMT and carotid plaque summary statistics were available from a meta-analysis of GWAS data in individuals of European ancestry for CIMT (up to 71,128 participants from 31 studies) and carotid plaque (up to 48,434 participants from 17 studies; 21,540 with defined carotid plaque).^23^

Genotyping and bioinformatic genetic analysis of each of the GWAS cited followed standardized procedures that are harmonized and comparable across the studies. Details are available in the studies cited.^16-20, 22, 23^ All the summary statistics were obtained from inverse-variance weighted meta-analysis restricted to participants of European ancestry after adjusting for age, sex, and principal components reflecting ancestry. eGFR and UACR were log-transformed. The meta-analysis in the BP GWAS additionally adjusted for BMI and genotyping platform.

### Statistical analysis

We performed summary-level univariable MR analyses to investigate the total, indirect, and direct effects of CKD (clinical diagnosis, eGFR, and UACR) on each of the stroke phenotypes (*Figure 1*). The total effect is defined as the net effect of genetically predicted CKD on stroke risk irrespective of mechanism. The indirect effect is defined as the effect of genetically predicted CKD on stroke risk that is mediated through hypertension. The indirect effect was calculated with the product of coefficients method,^24, 25^ in which we multiplied the univariable MR estimate for the effect of CKD on SBP and DBP and the multivariable MR estimate for the effect of SBP and DBP on stroke risk controlling for CKD. To test the null hypothesis of no mediation through hypertension, we calculated confidence intervals (CIs) for the indirect effect using the previously described Monte Carlo method.^26^ We used the propagation of error method to derive a P value for the indirect effect.^27^ We also calculated the proportion of the mediated effect by dividing the indirect effect by the total effect. The direct effect is defined as the association of genetically determined CKD on stroke risk through mechanisms independent of mediation. To estimate the direct effect, we used multivariable MR.^28^

For MR analyses, we used as instruments genetic variants that were associated with CKD diagnosis, eGFR, UACR, SBP, and DBP at genome-wide significance (p<5×10^−8^) after pruning based on linkage disequilibrium between variants (r^2^ <0.001). The instruments for kidney traits are presented in *eTable 1* and those used as proxies for BP traits have been previously published.^29^ The genetic association estimates between the instruments and the odds of the described outcomes were extracted from the MEGASTROKE and ISGC GWAS summary statistics. Following extraction of the association estimates and harmonization of the direction of the estimates across studies based on the effect allele, we calculated individual MR estimates for each instrument using the Wald estimator; standard errors were calculated using the delta method.^30^ We then pooled the individual MR estimates using fixed-effects inverse variance weighted (IVW) analyses. The overall effect sizes on stroke risk were reported as odds ratios (ORs) and 95% CIs of OR. Heterogeneity across estimates was examined with the I^2^ and the Cochran Q test (I^2^ > 50% and p<0.05 were considered statistically significant).^31^ We performed sensitivity analyses that are more robust than the IVW method to certain forms of pleiotropy, including the weighted median^32^ and MR-Egger.^33^ The estimated value of the intercept in MR-Egger regression can be interpreted as an estimate of the average pleiotropic effect across the genetic variants. An intercept term that differs from zero is indicative of overall directional pleiotropy.^33^ All MR analyses were performed in R (v4.1.0; The R Foundation for Statistical Computing) using the MendelianRandomization and TwoSampleMR packages. Finally, we also tested for the inverse association using kidney traits as outcomes and cerebrovascular disease phenotypes as exposures (bidirectional MR).^26^ Given the 6 cerebrovascular disease phenotypes studied for each of the 3 kidney traits, for the univariable MR analysis we set statistical significance at an FDR-adjusted p value < 0.05.

### Data availability

Genetic variants used are available in the supplemental information (*eSupplement*) and the code used for all analyses is available on request.

## RESULTS

### Univariable Analyses

There were 23, 195 and 38 genetic instruments for CKD diagnosis, eGFRcys and UACR, respectively. The genetic instruments explained 0.13% of the variance in CKD diagnosis, 1.2% of the variance in eGFRcys, and 0.16% of the variance in UACR, with mean F statistics of 23, 28, and 17, respectively.

Using fixed-effects IVW MR analysis, we found genetic predisposition to CKD diagnosis to be associated with a higher risk of AS and IS subtypes (OR, 1.07; 95% CI, 1.01-1.12; p=0.018 and OR, 1.07; 95% CI, 1.01-1.13; p=0.031, respectively) (e*Figure 1* and *eTable 2*). Furthermore, we also found genetically determined lower eGFR to be significantly associated with AS (OR per 1-log decrement in eGFR, 1.77; 95% CI, 1.31-2.40; p<0.001), IS (OR, 1.81; 95% CI, 1.31-2.51; p<0.001), and most strongly with LAS (OR, 3.00; 95% CI, 1.33-6.75; p=0.008) (e*Figure 1*). Associations between eGFR and these stroke subtypes remained significant even after FDR-adjustment of the P values (*eTable 3*). Genetically determined UACR was not significantly associated in univariable MR analysis with any of the cerebrovascular disease subtypes (e*Figure 1*). There was significant heterogeneity as defined by the Cochran Q test when exploring the associations between genetically determined eGFR, AS and IS, and between UACR and CES, but none of the intercepts from the MR-Egger regression were statistically significant, supporting a lack of significant pleiotropy in the analysis. Furthermore, the weighted median and the MR-Egger regression analyses provided association estimates that were directionally consistent and of similar magnitude as the ones derived from the IVW analyses, although with wider CIs, as expected given the lower statistical power of these approaches (*eTables 2-4*).

### Multivariable Analyses

We then performed MVMR controlling for SBP and found that there was still strong evidence for an independent effect of impaired kidney function on risk of ischaemic stroke (*Figure 2*). In particular, genetically predicted lower eGFR was still associated with risk of LAS even after controlling for SBP (OR per 1-log decrement in eGFR, 4.35; 95% CI, 1.84-10.27; p=0.001) (*Figure 2* and *eTable 3*). There was also strong evidence for an independent effect of genetically predicted lower eGFR on AS (OR, 1.98; 95% CI, 1.39-2.82; p<0.001) and IS (OR, 2.16; 95% CI, 1.48-3.17; p<0.001) subtypes. Genetic predisposition to CKD diagnosis also remained significantly associated with AS (OR, 1.07; 95% CI: 1.02-1.12; p=0.01) and IS (OR, 1.07; 95% CI: 1.01-1.12; p=0.01) subtypes after controlling for SBP in the MVMR analysis (*Figure 2* and *eTable 2*). As per the univariable analysis, the independent causal effects estimated from multivariable MR-Median and MR-Egger were consistent with the IVW analysis (*eTables 2-4*).

**Figure 2:**
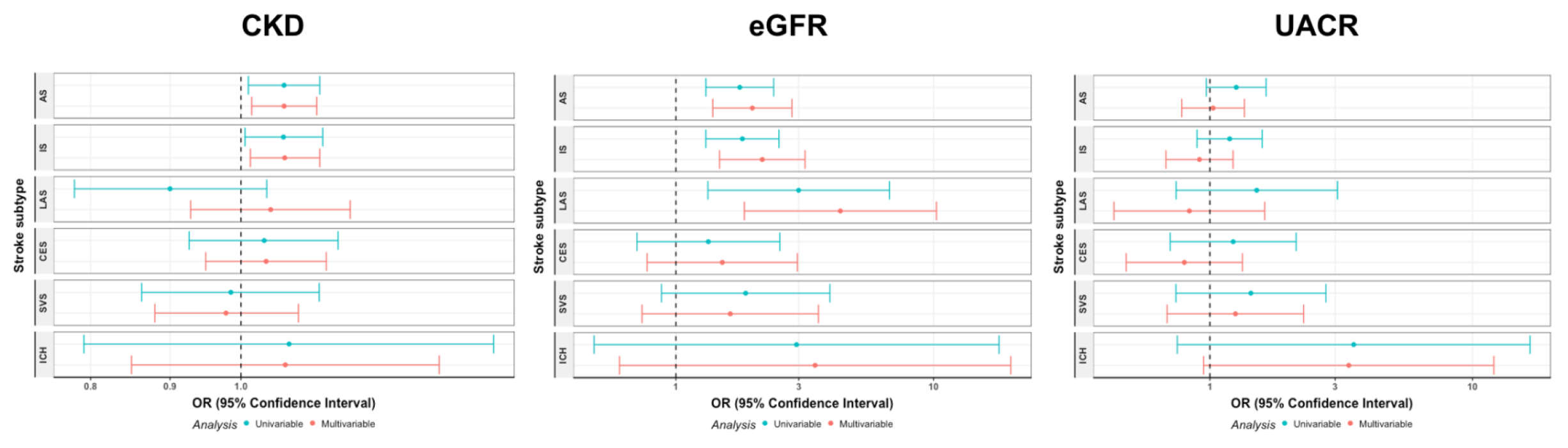
Univariable and multivariable Mendelian randomization associations between genetically determined kidney disease traits and cerebrovascular disease phenotypes, controlling for systolic blood pressure. Shown are the results derived from inverse variance weighted Mendelian randomization analysis. eGFR indicates estimated glomerular filtration rate; OR, odds ratio; UACR, urine albumin-to-creatinine ratio.

In multivariable MR analysis controlling for DBP, we found consistent evidence for a causal independent effect of genetically predicted CKD on AS subtype (OR, 1.06; 95% CI: 1.01-1.11; p=0.029) (*eTable 5* and *eFigure 2*) and genetically predicted eGFR on AS (OR per 1-log decrement in eGFR, 1.90; 95% CI, 1.33-2.72; p<0.001), IS (OR, 1.96; 95% CI: 1.33-2.90; p=0.001) and LAS (OR, 2.76; 95% CI: 1.16-6.58; p=0.02) (*eTable 6* and *eFigure 2*). Genetically determined UACR was also associated with ICH risk in the multivariable MR controlling for DBP (OR per 1-increment, 4.90; 95% CI: 1.38-17.46; p=0.014), but not SBP (*eTable 7* and *eFigure 2*).

### Mediation Analyses

We then performed mediation MR analysis in which we calculated the proportion of the effect that was mediated by BP by dividing the indirect effect by the total effect for each of the renal traits and cerebrovascular phenotypes. The most relevant mediation analysis results are presented in *Figure 3*. For the effect of genetically determined lower eGFR on odds of LAS mediated through SBP, we estimated an OR of 1.08 (95%CI: 1.01-1.14) and the calculated proportion mediated was 7.2% of the total effect (p=0.03). Similarly, for the indirect effect of lower eGFR on risk of LAS mediated through DBP was 1.09 (95% CI: 1.05-1.14) with DBP mediating only 7.8% of the total effect (p=0.01) (eFigure 3). For the genetic association between UACR and ICH risk, only 4.3% of the proportion of the total effect was also mediated by DBP (eFigure 3).

**Figure 3:**
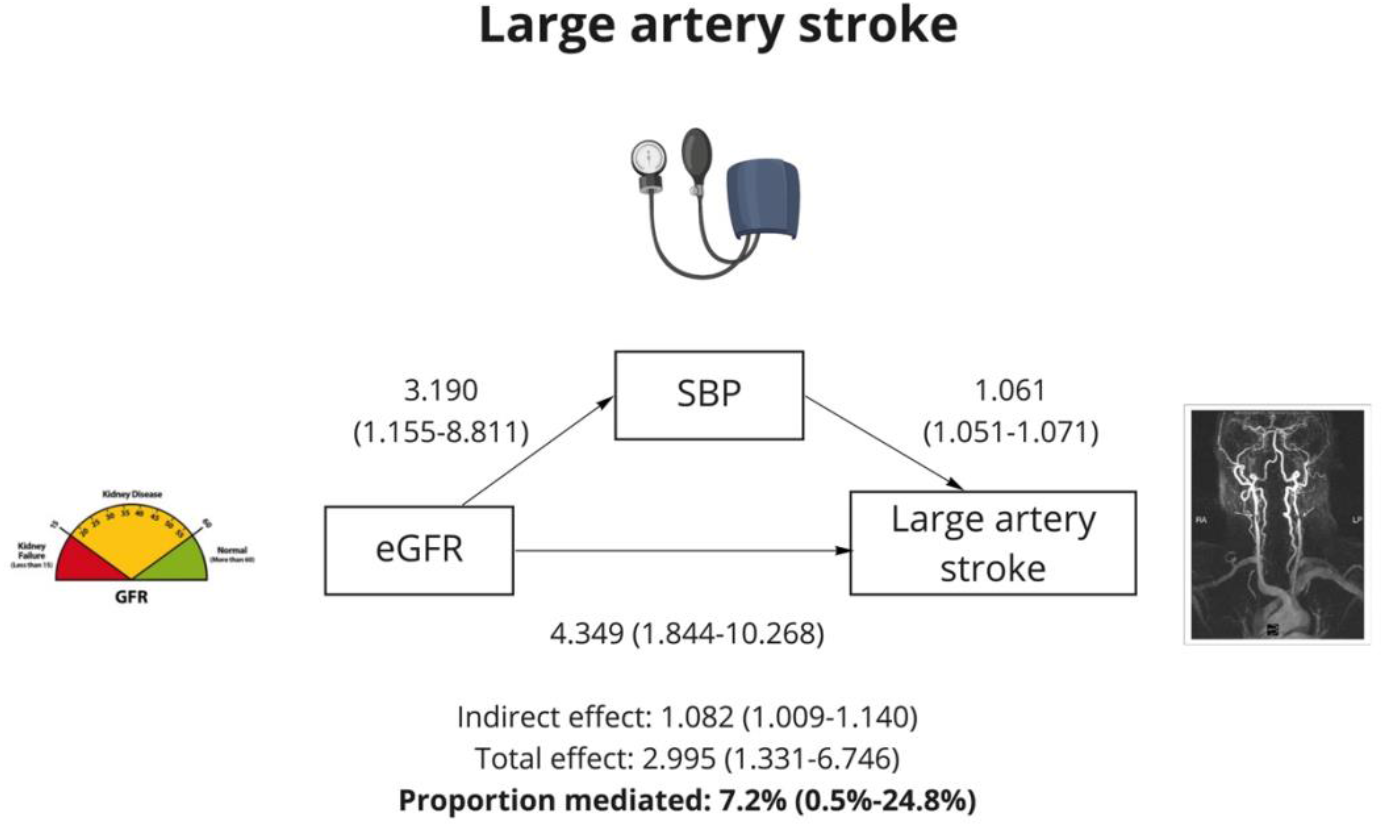
Mediation Mendelian randomization association between genetically determined estimated glomerular filtration rate and large artery stroke, controlling for systolic blood pressure. Directed acyclic graph demonstrating the total, direct and indirect effects of estimated glomerular filtration rate on large artery stroke risk as well as the proportion of the total effect mediated by systolic blood pressure. Shown are the results derived from inverse variance weighted Mendelian randomization analysis. eGFR indicates estimated glomerular filtration rate; SBP, systolic blood pressure.

To exclude reverse causation, we performed MVMR exploring the effects of BP traits on cerebrovascular phenotypes and what proportion of these effects were mediated by kidney disease traits (*eFigure 4*). There was no evidence of a bidirectional relationship between SBP and eGFR, and between DBP and eGFR with little or no proportion of the total effect of BP traits on stroke subtypes mediated by lower eGFR (<1%).

To explore the association between genetically predicted lower eGFR and LAS is mediated by effects of low eGFR on atherogenesis, we examined the relationship between lower eGFR and carotid intima media thickness as well as with presence of carotid plaque (eFigure 5 and *eTable 8*). However, we found no significant associations between lower eGFR and either carotid artery trait (Beta for CIMT, -0.004; 95% CI: -0.032-0.025; p=0.81, and OR per 1-log decrement in eGFR for carotid plaque, 0.98; 95% CI: 0.37-2.61; p=0.60).

## DISCUSSION

Leveraging large-scale genetic data, we applied univariable and multivariable MR to determine whether impaired kidney function influences risk of stroke independently of elevated BP. Little of this effect appeared to be mediated by BP. These results were largely robust to sensitivity analyses accounting for horizontal pleiotropy.

Our results build upon extant efforts to leverage genetics to explore potential causal relationships between kidney disease and stroke, with the important addition of MVMR controlling for BP traits. As such, we have found strong evidence for a direct causal effect of impaired kidney function on risk of stroke, particularly between genetically determined lower eGFR and LAS. By conducting mediation analysis in the context of MR, our study adds to knowledge by formally investigating whether increased BP mediates the effect of increasing stroke risk for patients with CKD. Our study provides evidence of little indirect effect of CKD on stroke risk through BP, suggesting that other mechanisms, possibly intrinsic to kidney disease, may underlie the pathophysiological processes leading to stroke in patients with CKD.

Deploying more recent and larger datasets,^16, 17^ we have also confirmed previous associations between genetically determined impaired kidney function and risk of IS demonstrated by both PRS and MR approaches.^10, 34^ In particular, consistent with the previous univariable MR study,^10^ there was a strong (3-fold) association between lower eGFR and LAS. Our results extend these previous findings by additionally showing associations between lower eGFR, AS, and IS, as well as between genetic predisposition to CKD diagnosis, AS and IS. The consistency of these relationships across various kidney disease traits and stroke subtypes provide further support for the cerebrorenal paradigm and for the potential causal role of CKD in cerebrovascular disease. In contrast to the previous MR study,^10^ we did not replicate the associations between genetic predisposition to higher UACR and LAS or ICH risk. However, this discrepancy likely relates to our stricter threshold for clumping (r^2^<0.001 vs <0.1 in the earlier study) as the findings were otherwise of similar magnitude and directionally consistent.

Although several large meta-analyses that have adjusted for traditional cardiovascular risk factors support our findings of an independent effect of impaired kidney function on stroke risk,^3, 8, 35^ our results do contrast with a more recent meta-analysis in which the association was attenuated with adjustment for measures of longitudinal BP control.^7^ These conflicting results may relate to lack of statistical power or subtype specificity, study heterogeneity, and the inherent limitations of pooling mostly observational studies for meta-analysis. In contrast, our study benefits from the statistical power of the largest and newest GWAS datasets available including the phenotypic characterization of stroke into its etiopathologic subtypes.

Several mechanisms have been suggested to explain the observed associations between CKD and stroke.^4, 5^ One might have expected a causal link between CKD and SVS, but if hypertension mediates both of these then it could account for the lack of causal association. ^36^ There was instead a stronger, independent association with LAS, implicating an impact of kidney disease on the atherosclerotic disease pathways in some way. The absence of a corresponding association with CIMT suggests that CKD may not necessarily affect atherogenesis but rather that it may play a role in atheroprogression, plaque destabilization or rupture. In observational studies, patients with CKD have been shown to have different plaque morphology with reduced total collagenous fibre content, greater levels of total calcification, and higher lesion stability and risk of rupture.^37^ Furthermore, serum markers of inflammation, vascular calcification, and vessel wall degradation have also been shown to be significantly elevated in CKD patients compared to those with normal renal function including enhanced levels of fibrinogen, parathyroid hormone, fetuin-A, and matrix metalloproteinase-7, suggesting that the proinflammatory milieu of CKD may play a role.^38^ However, further studies are required to better understand the impact of CKD on LAS pathophysiology.

Our study had a number of strengths. Compared to conventional observational studies, MR is more robust to residual confounding and measurement error since genetic variants are predisposed, randomly assigned at birth and are thus often less confounded indicators of particular traits.^39^ This current study applied both univariable and multivariable MR methods, using the largest number of SNPs identified from the latest well-powered GWAS studies that could be identified in the literature. MVMR is a novel extension of MR analyses that allows exploration of causality and clarification of the direction of demonstrated associations, thus ruling out reverse causation.^39^ A series of pleiotropy-robust methods were rigorously applied to explore the possibility that findings were not biased as a result of pleiotropy.

Our study also has some limitations. Firstly, using instruments with an F-statistic > 10 only reduces bias to less than a certain level, and problems with weak instrument bias still occur.^40^ Secondly, as there was less statistical power in the smaller sample of ICH, we may not have captured all potential causal associations between impaired kidney function and ICH. Thirdly, we cannot exclude the possibility of confounding by cryptic pleiotropy, which is an established limitation of MR analyses.^41^ However, our multimodal approach including methods for quantifying pleiotropy and multiple MR approaches with different modelling assumptions regarding the use of pleiotropic variants in the analyses provides some reassurance for the validity of our MR models. Fourthly, analyses were restricted to individuals of European ancestry, which may limit the generalisability of our results to other ancestry groups. This is particularly relevant to vulnerable populations such as Black and Hispanic patients where there are important phenotypic and prognostic differences in hypertension and CKD.^42, 43^ One previous trans-ancestry MR analysis did not demonstrate an association between eGFR and stroke risk; however, this study focused on all ischaemic stroke and not individual cerebrovascular phenotypes, and they did not perform mediation analysis in the context of MR.^44^

In conclusion, this study used both univariable and multivariable MR analyses to demonstrate a causal effect for impaired kidney function on stroke risk that is independent of hypertension. In particular, we observed large effects for LAS risk in our multivariable MR analysis. These findings could help prioritize CKD prevention and treatment to mitigate stroke risk and motivate future studies to provide further insight into mechanisms linking kidney disease and stroke.

## Supporting information

eSupplement

## Data Availability

Genetic variants used are available in the supplemental information (eSupplement) and the code used for all analyses is available on request.

